# Exploring the Correlation Between the COVID-19 Pandemic and Increased Daily Cigarette Consumption in Yogyakarta, Indonesia: A Machine Learning Approach

**DOI:** 10.1101/2023.09.30.23296376

**Authors:** Desy Nuryunarsih, Lucky Herawati, Atik Badi’ah, Jenita doli tine donsu

## Abstract

**Objective:** Smoking is very common in Indonesia: among adults, around 66% of males and 7% of females are smokers. Smoking is not only harmful for people who smoke but also for people who are exposed to second-hand smoke on a regular basis. Previous research in various countries has shown a changing trend in smoking during the COVID-19 pandemic. However, despite the high prevalence of smoking in Indonesia and the shifting trend during COVID-19, no studies have utilized machine learning to investigate the potential increase in daily cigarette consumption during the pandemic. This study aimed to predict the increase in daily cigarette consumption among smokers during the pandemic, focused on smokers selected from vaccination registrants in the Special Region of Yogyakarta.

**Design:** Five machine learning algorithms were developed and tested to assess their performance: decision tree (DT), random forest (RF), logistic regression (LoR), k-nearest neighbors (KNN), and naive Bayes (NB). The results showed a significant difference in the number of cigarettes consumed daily before and during the pandemic (statistic=2.8, p=0.004).

**Setting:** This study is believed to be the first study prediction model to predict the increase of cigarette consumption during the COVID-19 pandemic in Indonesia.

**Results:** The study found that both DT and LoR algorithms were effective in predicting increased daily cigarette consumption during the COVID-19 pandemic. They outperformed the other three algorithms in terms of precision, recall, accuracy, F1-score, sensitivity, and AUC (area under the curve operating characteristic curve). LoR showed a precision of 92%, recall of 99%, accuracy of 93%, F1-score of 96%, sensitivity of 91% and AUC of 78%, DT showed a precision of 88%, recall of 91%, accuracy of 81%, F1-score of 89%, sensitivity of 95% and AUC of 98%.

**Conclusion:** We recommend using the DT and LoR algorithms, as they demonstrated better prediction performance. This study can be used as a pilot study for predicting smokers’ continuing behaviour status and the possibility of smoking cessation promotion among smokers, this study is a short report, and we suggested expanding with more factors and a larger dataset to provide more informative and reliable results, The recommendations based on the current findings can serve as a starting point for initial actions and can be further validated and refined with larger-scale studies in the future.

**STRENGHTS AND LIMITATION OF THIS STUDY:**

⟹ This is the first study to investigate the increased number of cigarettes consumed daily by Indonesian smokers during the pandemic using machine learning models.
⟹ This paper using Multiple Algorithms: The author did not rely on a single algorithm but compared five different ML methods, providing a comprehensive analysis.
⟹ This paper using external research as a reference, the author established a solid basis for their methodology and ensured their research was supported by existing literature.
⟹ The paper clearly identified the DT model as superior, bringing clarity to the readers.
⟹ The paper suggests that the developed framework has wide applicability in healthcare, increasing its relevance and potential impact.
⟹ This paper considered only a few features (27), and more data on economic factors can be incorporated in future research work, as it will enable the real-life application of this model.
⟹ The selection bias introduced by recruiting participants from those who came for vaccination. This sample may not fully represent the general population.

## Introduction

Smoking is very common in Indonesia: among adults, around 66% of males and 7% of females are smokers. Smoking is not only harmful for people who smoke but also for people who are exposed to second-hand smoke on a regular basis. An estimated 90 million Indonesian people involuntarily inhale cigarette smoke every day. Every hour 46 Indonesians die due to smoking-related diseases [1]. Around 90% of Indonesian smokers use Kretek, this clove cigarettes carry similar health risks as regular cigarettes, recent evidence highlights the comparable harm of kretek cigarettes [2]. During the COVID-19 pandemic, there have been changes in smoking behaviour. Some individuals have reported smoking more due to stress or anxiety, while others have used this time as an opportunity to reduce smoking [3].

Smoking is commonly perceived, particularly among smokers, as a way to regulate emotions and relieve negative emotions [4]. Interestingly, although COVID-19 has indeed caused significant stress for many people, a study conducted in Hong Kong showed that overall tobacco use decreased after the first two waves of the COVID-19 outbreak in Hong Kong. A greater proportion of cigarette users decreased use compared to that of HTP and e-cigarette users [5]. Similarly, in Japan, studies suggest that during the COVID-19 state of emergency people who have high-risk factors for COVID-19 might change their smoking behaviour for the better, while people who work from home or live alone might change their smoking behaviour for the worse [3].

Furthermore, COVID-19 lockdown policies and self-isolation forced people to stay at home worldwide, and schools were temporarily closed during the pandemic in most countries. Smokers who usually spend more hours outdoors, such as smoking in the park, workplace, or school, may consequently spend more time smoking indoors [6]. Studies show that the COVID-19 lockdowns had wide-ranging economic impacts [7]. Many people lost their jobs or had salary reductions, which added to the negative health impacts of lockdowns themselves, reflected in increased sedentary behaviours, loneliness, isolation, anxiety, depression, and stress [8].

Certain factors are associated with increased smoking during the COVID-19 pandemic such as lower education level, living alone, poor subjective health status and anxiety about economic damage due to COVID-19, Furthermore, study in Italy showed that the COVID-19 lockdown had a significant impact on smoking consumption. People experiencing a decline in quality of life, reduced sleep, increased anxiety, and depressive symptoms were more likely to report increased cigarette consumption (OR: 2.05; 95% CI: 1.49 to 2.80 for quality of life, OR: 2.29; 95% CI: 1.71 to 3.07 for sleep, OR: 1.83; 95% CI: 1.38 to 2.43 for anxiety, and OR: 2.04; 95% CI: 1.54 to 2.71 for depressive symptoms) [9].

By utilizing statistical models and advanced techniques like machine learning, researchers can analyse complex data to predict the increase in daily cigarette consumption among smokers during the pandemic and uncover hidden patterns. This research provides valuable insights into smoking behaviour during challenging times. Predictive modelling, a sophisticated approach that employs advanced mathematics and computational techniques, enables researchers to forecast future events or outcomes. It has great potential for forecasting what will happen in the future.

When predictive modelling is implemented effectively, it has shown remarkable outcomes in different medical fields. It can help forecast disease progression and identify high-risk patients who need early intervention. There are numerous successful examples of using this approach in healthcare settings worldwide [10-12]. The challenge of predicting cigarette consumption before and during the pandemic using traditional statistics is that it relies on predefined assumptions and models, these methods may not fully capture the complex and dynamic nature of smoking behaviour. On the other hand, using machine learning allows for more flexibility and adaptability in analysing data, identifying patterns, and making accurate predictions. Machine learning can uncover hidden insights and provide a more nuanced understanding of smoking behaviour during different periods. It is a powerful tool for enhancing prediction accuracy. Our study uses machine learning (ML) techniques to analyse and predict changes in smoking behaviour. ML, which involves the study and development of algorithms that learn from data, allows for a more comprehensive understanding of the impact of the pandemic on cigarette consumption [13, 14].

Once we have a comprehensive understanding of the impact of the pandemic on cigarette consumption through ML techniques, we can use this knowledge to develop targeted interventions and strategies to promote healthier behaviours. This study can lead to a valuable overview of how data-driven insights can be used to make a positive impact.

In this study, we conducted a cross-sectional survey focusing on smokers from the Special Region of Yogyakarta, Indonesia. Our goal was to predict the rise in daily cigarette consumption among smokers during the pandemic. We considered various factors related to smoking and COVID-19 and we developed and tested five machine-learning algorithms: decision tree (DT), random forest (RF), logistic regression (LoR), k-nearest neighbors (KNN), and naive Bayes (NB), we evaluated their accuracy in predicting the increase in daily cigarette consumption during the pandemic. By incorporating these diverse machine learning techniques, we were incorporating the latest ideas in the field. Each approach has its unique strengths that contribute to a comprehensive and robust predictive model. In the methodology section, we will provide a detailed explanation for each model.

## Objectives

This study aims to use ML algorithms to predict the increase in cigarette consumption among participants during the COVID-19 pandemic. We include various features that influence smoking status during COVID-19, such as pre-pandemic smoking status, stress levels, concerns about getting infected, presence of systemic diseases, anxiety related to COVID-19 complications and death, and health insurance status. In the future, this predictive model could be valuable for evaluating post-COVID-19 cigarette consumption.

## Materials and methods

This study used a cross-sectional survey with 131 respondents randomly selected from vaccination registrants in the Special Region of Yogyakarta, from August to October 2021. They filled out the questionnaire voluntarily after receiving the vaccination. The questionnaire contained general demographic items, covering age, gender, home location, education, and occupation, and it asked questions about health status, experiences with negative emotions during COVID-19, smoking status, amount of tobacco consumption, changes in smoking behaviour during the COVID-19 pandemic, and any economic and social factors affecting smoking to the pandemic.

The datasets were classified or grouped using machine learning into input and output features. The input features (X) and output (y) are explained above in detail. The output variable, called the target feature, was binomial data. The model prediction algorithms used were ANN, LoR, NB, DT and RF.

Artificial neural networks (ANN) are powerful algorithms that employ the principles of probability to create a classification prediction model. By utilizing data on past events, the model can predict what may occur in the future. The ANN are inspired by the structure of neurons in the mammalian cortex. They mimic the way our brain works to process information and make predictions. ANN calculates the probability of an event and can adapt if additional supporting information is provided. Despite its impressive capabilities, ANN employs a black-box approach, meaning that it is difficult to discern the internal processes that are responsible for generating predictions.[12, 15] Logistic Regression (LoR) is a type of regression that uses binary categories instead of numeric ones. In developing a logistic regression model, we use observation features and their corresponding labels to train the model. Once trained, the model can make predictions for new data. It is commonly used for binary classification, specifically predicting whether an event will occur or not [16]. The Naive Bayes (NB) algorithm is a set of classification techniques developed based on Bayes’ theory [17]. The primary function of NB is to calculate the probability of an observation being categorized into a particular group (class). This is achieved by using a training dataset to compute the probability of each class based on its features. When the model is faced with new data, it applies the new features to calculate the likelihood of each class being the correct one. In statistics, Bayes’ theory explains the concept of conditional probability [16], which is the probability of event A occurring given that event B has taken place. Event A is dependent on event B; hence this is referred to as conditional probability. A decision tree is a machine learning algorithm that employs a set of rules arranged in a tree-like structure to make decisions by modelling potential outcomes. This algorithm utilizes data attributes to divide data into smaller groups, which are repeatedly split until all data elements belonging to the same class fit into one group [16]. The performance of the models used in this study was assessed through the calculation of various metrics, including percentage accuracy, precision, F-1 score, and sensitivity. The accuracy and precision of the models were evaluated by measuring the amount of data correctly predicted as positive, relative to all data predicted positively, including both true and false positives. Specifically, the study involved making predictions based on 27 features to determine whether the participants increased smokers’ cigarette consumption daily.

The sensitivity or recall of these model algorithms was also measured, indicating the number of instances that were correctly predicted as positive relative to all positive data. This metric provides insight into how many instances the models missed in predicting the increase of participants’ cigarette consumption who should have been identified as such. Additionally, the F-1 score was calculated as the harmonic mean of precision and recall, with the best score being 1.0 and the worst value being 0. A high F-1 score indicates that the classification model has good precision and recall, thereby representing an accurate and reliable model. We were also looking into the area under the curve operating characteristic curve (AUC) to evaluate the performance of our binary output, the AUC represents the ability of a model included to distinguish between positive and negative instances, with an AUC value of 1 indicating model makes better predictions and less than 0.5 indicates that a model performed no better than random chance.

### Data preparation

The primary data from 131 participants’ survey results were filtered, and text variables were transformed into numerical, scaled datasets and normalized before correlation statistics were performed. There were three missing data in the age feature, we used an imputation technique using the median value of the participant’s age, we found one outlier in each number of cigarettes consumed before the pandemic and after the pandemic, and we did an imputation using normal data range with lower limit as Q1-1.5*IQR and upper limit as Q3+1.5*IQR. Finally, recursive feature elimination (RFE) was performed to determine the rank of each feature or variable that produced the best prediction model.

### Cigarette consumption before and during the pandemic

Before further data analysis, the features influencing increase cigarette consumption were determined. A paired t-test was performed to determine whether there was a difference between the number of cigarettes consumed daily before and during the pandemic.

### Machine learning algorithms

To compare machine learning algorithms, the study population was split into a ‘training’ group, in which the features included in algorithms were derived, and a ‘test’ dataset. The ‘training’ dataset was derived from a random sampling of 80% of the extracted data set and the validation set data comprised the remaining 20%.

### Analysis

In this study, we utilised statistical analysis and ML models. A t-test analysis to show the difference between smoking before and during the pandemic, analysis then followed ANN (Artificial Neural Network), LoR (Linear regression), NB (Naïve Bayes), DT (Decision Tree) and Random Forest (RF).

Models were developed with Python 3.7 (Python Software Foundation, Wilmington, DE, USA). For the initial analysis, we used a paired t-test scipy stats. The ANN was designed using MLP Classifier (hidden layer size= 3, maximum iter = 5). With a limited amount of data, using a hidden layer size of 3 and a maximum iteration of 5 can help prevent overfitting and keep the model from becoming too complex. We conducted several exercises before deciding on these specific hyperparameters to strike the right balance between model complexity and the available data. It is important to find the best combination that works for our study. For the other methods, the training process was achieved using scikit-learn 0.21.2.

### Ethical approval

The study was approved by the ethic committee of the Polytechnic of Health Department Yogyakarta, Republic of Indonesia, with the approval number No. e-KEPK/POLKESYO/0751/X/2021, granted on October 5th, 2021.

## Results

### Participant characteristics

From the 131 respondents, 128(97.7%) were male, and 3(2.2%) were female. Before the pandemic, a total of 108(82.4%), 13(9.9%), and 10(7.6%) participants smoked kretek, white, or any type of cigarettes, respectively, similarly. During the pandemic, 107(81.6%), 14(10.6%), and 10(7.6%) participants smoked kretek, white, or any type of cigarettes, respectively. Participants’ age distribution ranged from 18 to 70 years, with a mean of 35.3 ±12.4 (Table 2)

**Table 1.** Selected features and information.

| Feature | Type | Information |
| --- | --- | --- |
| Sex | Nominal | Gender category:<br>Male<br>Female |
| Education | Nominal | Education<br>Primary school<br>Junior high school<br>High school<br>Vocational<br>Bachelor<br>Master<br>PhD |
| Meet people | Nominal | Whether participants regularly meet people during the pandemic<br>Yes<br>No |
| Stress | Nominal | Whether or not participants feel stress at least once a day during the pandemic.<br>Yes<br>No |
| Nervous | Nominal | Whether or not participants feel nervous at least once a day during the pandemic.<br>Yes<br>No |
| Disease | Nominal | Whether or not the participant has at least one of the diseases: lung problem, diabetes, heart disease, allergy<br>Yes<br>No |
| Insurance | Nominal | Whether or not the participant has insurance<br>Yes<br>No |
| Product before | Nominal | What type of cigarettes did participants smoke daily before the pandemic?<br>Kretek<br>White cigarette<br>Both |
| Amount before | Numeric | How many cigarettes were consumed daily before the pandemic (cigs) |
| Important smoking before | Nominal | Do you think it is important to smoke cigarettes (before the pandemic)<br>Yes<br>No |
| Product pandemic | Nominal | What type of cigarettes participant smokes daily during the pandemic<br>Kretek<br>White cigarette<br>Both |
| Amount pandemic | Numeric | How many cigarettes were consumed daily in the pandemic (cigs) |
| Important smoking pandemic | Nominal | Whether or not participants think it is important to smoke a cigarette during the pandemic?<br>Yes<br>No |
| Smoking in | Nominal | Whether or not participants smoke cigarettes inside their house<br>Yes<br>No |
| Last smoking | Nominal | Whether or not participants smoke cigarettes after waking up in the morning<br>Yes<br>No<br>Don't remember |
| Total cigarettes daily | Numeric | The total number of cigarettes consumed daily is more than 25 cigarettes. |
| Smoking pandemic | Nominal | Whether or not participant smoke during the pandemic<br>Yes<br>No |
| Getting cigarettes | Nominal | Whether or not it is difficult to get cigarettes during the pandemic<br>Yes<br>No |
| Important stop | Nominal | Whether or not they think it is important to stop smoking cigarettes during the pandemic<br>Yes<br>No |
| Try to stop | Nominal | Whether or not they try to stop smoking during the pandemic<br>Yes<br>No |
| Change brand | Nominal | Whether or not they change their cigarettes type during the pandemic<br>Yes<br>No |
| Afraid get covid | Nominal | Whether or not they afraid of COVID-19 effect as a smoker<br>Yes<br>No |
| Complication | Nominal | Whether or not they think of the complication effect of COVID-19 and smokers<br>Yes<br>No |
| HCW intervention | Nominal | Whether or not they get intervention from HCW to quit smoking during the pandemic<br>Yes<br>No |
| Vaccine | Nominal | Whether or not they have gotten a vaccine for SARS-CoV-2<br>Yes<br>No |
| Vaccine smoking behaviour | Nominal | Whether or not they think it is important to get the vaccine as a smoker during the pandemic<br>Yes<br>No |
| Age group | Nominal | Age of participant<br>18-30<br>31-40<br>41-50<br>>50 |
| Cigarettes increase | Nominal | Whether or not cigarettes increased during the pandemic<br>Yes<br>No |

**Table 2.** Characteristic of participants.

| Variable | Frequency | Percentage |
| --- | --- | --- |
| Sex |  |  |
| Male | 128 | 97.7 |
| Female | 3 | 2.2 |
| Education |  |  |
| Primary school | 10 | 7.6 |
| Junior high school | 0 | 0 |
| High school | 61 | 46.0 |
| Diploma 3 | 0 | 0 |
| Bachelor | 28 | 21.3 |
| Master | 2 | 1.5 |
| PhD | 1 | 0.7 |
| Age group |  |  |
| 18-30 | 33 | 25.1 |
| 31-40 | 48 | 36.6 |
| 41-50 | 26 | 19.8 |
| >50 | 24 | 18.3 |
| Smoking products before pandemic |  |  |
| Kretek | 108 | 82.4 |
| White cigarettes | 13 | 9.9 |
| Any type of cigarettes | 10 | 7.6 |
| Smoking products during pandemic |  |  |
| Kretek | 107 | 81.6 |
| White cigarettes | 14 | 10.6 |
| Any type of cigarettes | 10 | 7.6 |

The paired t-test analysis revealed a significant difference (statistic=2.8, p-value=0.004) in the mean number of cigarettes consumed daily before and during the pandemic. This supports the alternative hypothesis that there was a slight increase in daily cigarette consumption during the pandemic (mean=7.5, SD=4.3 vs mean=8.6, SD=4.7) (Table 3).

**Table 3.** T-test analysis number of cigarettes before and during the pandemic.

| Number of cigarettes daily summary | Before pandemic | During pandemic |
| --- | --- | --- |
| count | 131 | 131 |
| mean | 7.5 | 8.6 |
| std | 4.3 | 4.8 |
| min | 1.0 | 0 |
| 25% | 5 | 5 |
| 50% | 7 | 7 |
| 75% | 10 | 12 |
| max | 20 | 20 |

Test\_relResult(statistic=array([2.8708006]), array([0.0047815a])
T-test p-value 0.004a a significant 0.005

The violin graph (Graph 1) below compares the number of cigarettes before and during the pandemic. It shows the impact of the pandemic on smoking habits. The x-axis represents the period, before and during the COVID-19 pandemic. The y-axis represents the number of cigarettes. The graph visually demonstrates the slight difference in smoking behaviour before and during the pandemic.

**Graph1.**
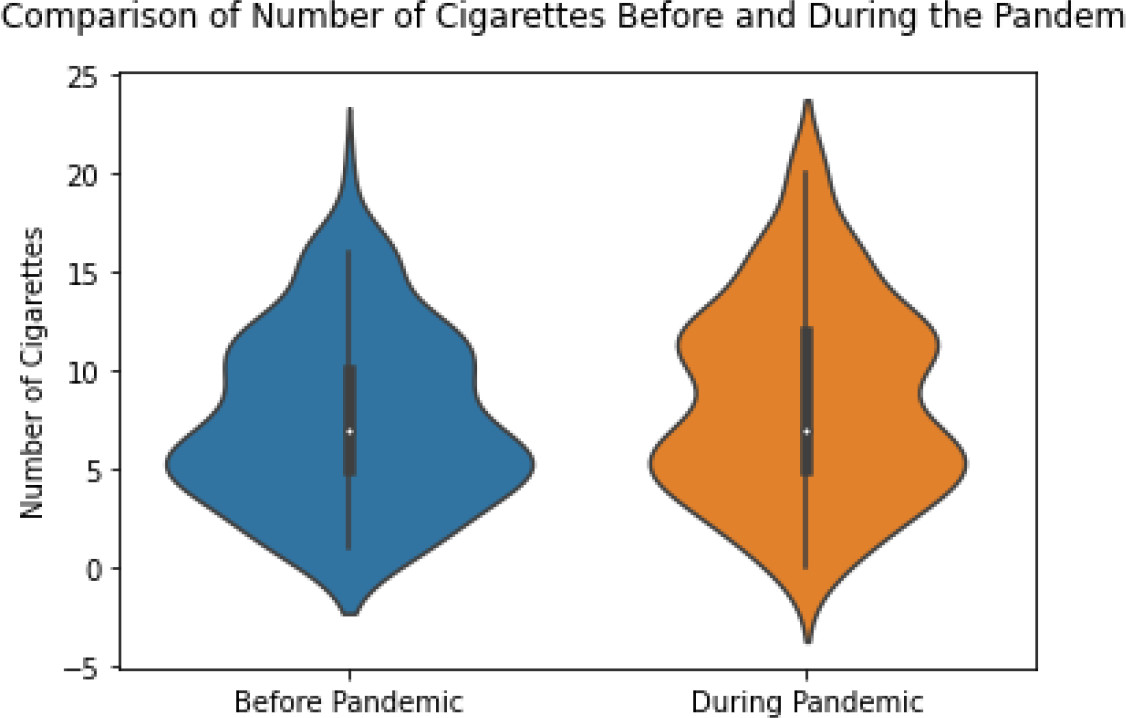
The number of cigarettes consumed daily before and during the pandemic.

### Feature selection and percent correlation of the feature to predict an increased number of cigarettes consumed

The graph below (Graph 2) displays the Pearson correlation matrix of the features, with different correlation scores represented by colour codes. The orange colour indicates a strong positive correlation, while the dark blue colour represents a strong negative correlation. The features that are most correlated with the number of cigarettes consumed during the pandemic will be further explained in Table 4.

**Table 4.** Feature selection and Pearson correlation of predictive factors for increased daily cigarette consumption.

| Variable | Pearson correlation | Feature selected rank |
| --- | --- | --- |
| Amount pandemic | -0.25 | 1 |
| Education | 0.20 | 2 |
| Try to stop | -0.18 | 3 |
| Last smoking | 0.17 | 4 |
| Smoking in | -0.15 | 5 |
| Nervous | -0.14 | 6 |
| Age | -0.13 | 7 |
| Vaccine | 0.13 | 8 |
| HCW_intervention | 0.12 | 9 |
| Amount before | 0.11 | 10 |
| Afraid get COVID-19 | 0.10 | 11 |
| Important smoking pandemic | -0.10 | 12 |
| Getting cigarettes | 0.09 | 13 |
| Total cigarettes daily | -0.09 | 14 |
| Insurance | 0.07 | 15 |
| Disease | 0.06 | 16 |
| Product pandemic | -0.06 | 17 |
| sex | -0.06 | 18 |
| Vaccine smoking behav | 0.06 | 19 |
| Smoking pandemic | 0.05 | 20 |
| Complication | 0.05 | 21 |
| Product before | -0.05 | 22 |
| Stress | -0.04 | 23 |
| Important smoking before | -0.02 | 24 |
| Meet people | -0.02 | 25 |
| Change brand | 0.02 | 26 |
| Important stop | 0.02 | 27 |

**Graph 2.**
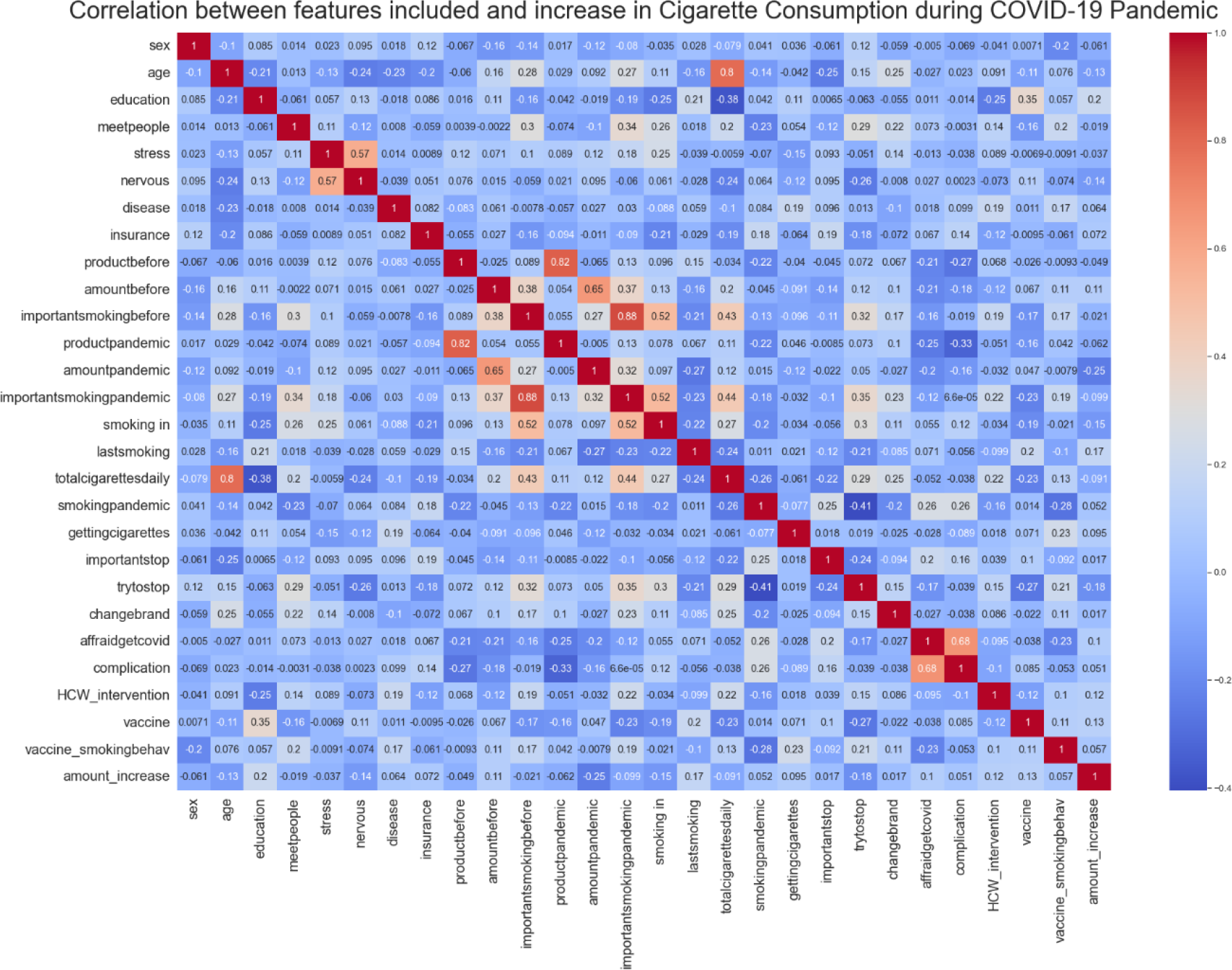
Pearson correlation matrix for the selected features

The table below presents the features that show the strongest correlation with the increase in the number of cigarettes smoked daily during the pandemic. The feature that shows the highest correlation is “Amount pandemic,” followed by “Education.” On the other hand, the features “Important stop” and “Changed brand” show the least correlation with the increase in cigarette consumption.

From the features obtained from the 131 randomized participants, the ten factors most correlated in order with an increase of cigarettes consumed during the pandemic were amount pandemic, education, try to stop, last smoking, nervous, age, vaccine, HCW_intervention, amount before, afraid get COVID-19. Information about these features selected can be found in (Table 4)

Model Performance after training the machine learning models with the training dataset. The sensitivity, precision, accuracy, F1-score and area under the receiver operating characteristic curve (AUC) show strong performance, for DT were 95%, 88%, 81%,89% and 84%. For LoR was 91%, 92%, 93% and 78%, respectively. Thus, we suggested adopting a decision tree and logistic regression algorithms, which achieved a better performance (Table 6)

**Table 6.** Model performance for increased smoking cigarettes consumed daily during the pandemic.

| Name | Precision | recall | Accuracy | F1-score | Sensitivity |
| --- | --- | --- | --- | --- | --- |
| ANN | 88 | 61 | 59 | 72 | 35 |
| LoR | 92 | 99 | 93 | 96 | 91 |
| NB | 23 | 22 | 33 | 36 | 50 |
| DT | 88 | 91 | 81 | 89 | 95 |
| RF | 88 | 91 | 81 | 89 | 85 |

The figure below shows AUC values that represent the performance of different classifiers. A higher AUC indicates better predictive performance. In this case, the decision tree has the highest AUC of 0.96, followed by logistic regression and random forest with AUCs of 0.78. Naive Bayes has the lowest AUC of 0.64. This means that the decision tree classifier is performing the best in terms of predicting the target variable, while Naive Bayes is performing the least accurately.

**Figure 2.**
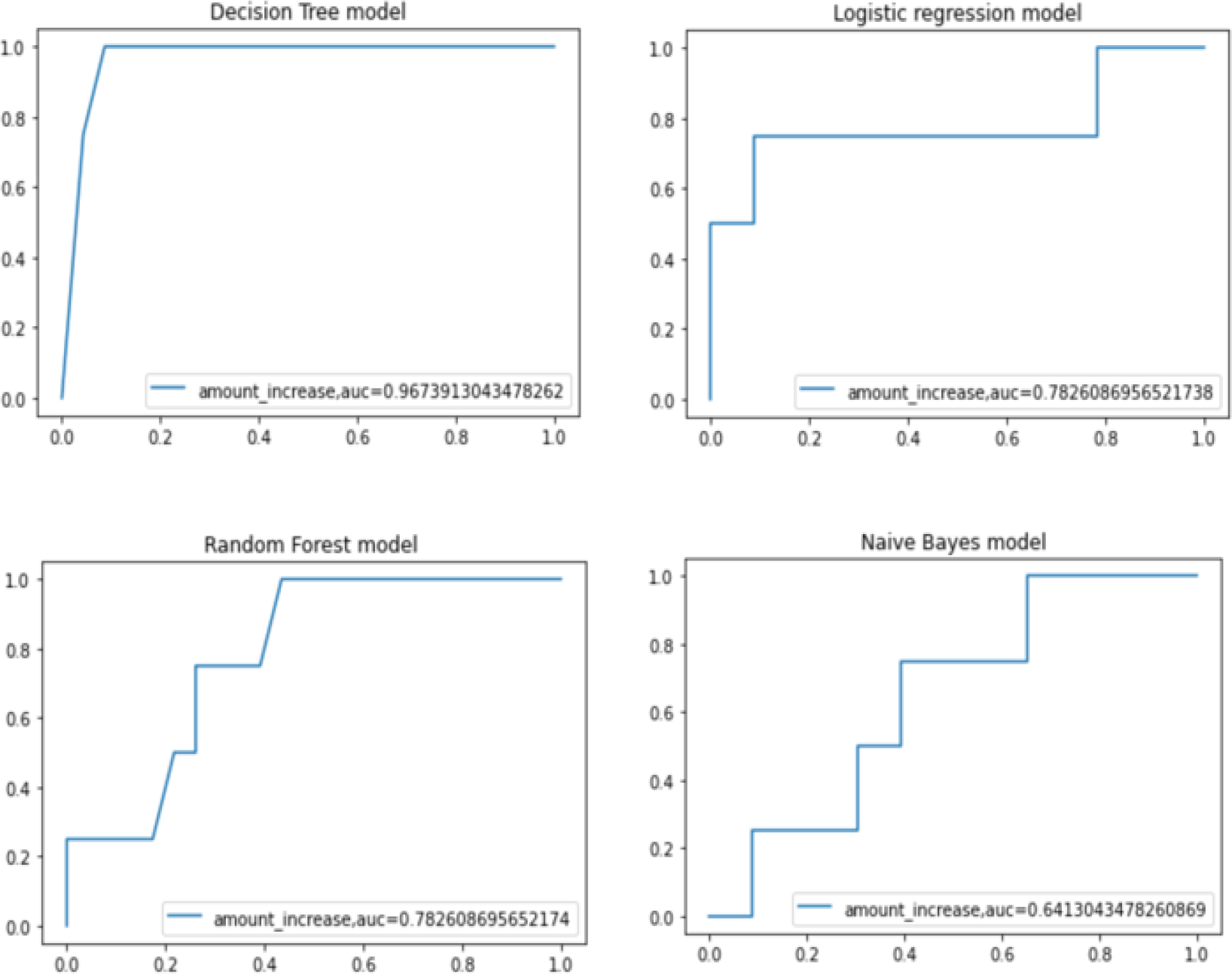
shows the under-the-curve area (AUC) c-statistic; a value close to 1 means the model is close to perfect while an AUC c-statistic below 0.5 means not a good model.

## Discussion

In this study, we compared the performance of different machine learning analysis techniques, including ANN, LoR, NB, DT, and RF. We evaluated their precision, recall, accuracy, F1 score, and sensitivity. When predicting smoking behaviour during the COVID-19 pandemic, we consider features that have been evaluated by other researchers and shown to influence smoking patterns. The experiments were carried out to identify an optimal approach for real-time prediction of an increased number of cigarettes consumed daily, using supervised ML-based multi-class classification methods.

Our objectives for this study were to compare different classification algorithms and identify the best classification method for predicting participant status of increased cigarette consumption during the COVID-19 pandemic, and to identify the most important features by conducting a feature selection analysis.

To develop an accurate classification model, the quality of the data on which it is to be trained becomes very important; in this study, we trained 80% of our data. To ensure the best quality data, data on factors influencing smoking status during the pandemic that have been investigated in previous research were included [3, 5, 9, 18]. Additionally, we include the insurance participant status as an additional factor in our predictive model, The relevance of “having health insurance” in predicting increased cigarette consumption during the pandemic might not be immediately apparent. However, it could be considered as a potential influencing factor. By exploring the relationship between health insurance coverage and health-related behaviours, including smoking habits, we can better understand the complex dynamics at play. It is important to consider various factors when predicting cigarette consumption during the pandemic to develop a comprehensive and robust predictive model. Previous works on this topic focused mainly on smoking cessation prediction for white cigarette use, mostly prediction about smoking cessation results.

The statistical analysis showed a significant increase in cigarette consumption during the pandemic. However, it is important to consider the practical significance of this finding. While the increase may be statistically significant, it is necessary to evaluate its real-world implications for public health. This involves considering factors such as the magnitude of the increase, the overall smoking patterns, and the potential impact on smoking-related health issues. By contextualizing the statistical significance within the practical context, we can provide a more comprehensive understanding of the findings. Additionally, conducting an ablation study helped validate the contribution and effectiveness of different machine-learning techniques in predicting increased cigarette consumption. This approach involved evaluating the influence of various features, including participant smoking status and health insurance, along with other factors identified by previous research.

In terms of analysis performance, the results showed that LoR had the highest precision and recall values, indicating its ability to accurately predict increased cigarette consumption. ANN and DT also performed well in terms of precision and recall. However, NB had lower precision, accuracy, and sensitivity values compared to the other techniques. RF showed consistent performance across different metrics. By conducting this study, we were able to validate the effectiveness of each technique and understand their contributions to the overall predictive model. These findings highlight the importance of considering different machine-learning approaches when predicting increased cigarette consumption. Naive Bayes (NB) showed lower precision, accuracy, and sensitivity compared to the other techniques because it assumes independence between features, which may not hold in the case of predicting increased cigarette consumption. This assumption can lead to limitations in capturing complex relationships and dependencies in the data, resulting in lower performance compared to other techniques like logistic regression (LoR), artificial neural networks (ANN), and decision trees (DT)

The study showed that DT and LoR have better performance compared to the other three algorithm models. DT approach resembles the human thought process, where humans rely on their experiences to make decisions [16]. The decision tree algorithm breaks down data by calculating information gain or using input information in the form of features, in this study we input 27 features in connection with smoking behaviour and the COVID-19 pandemic, while LoR was used to predict the probability of an event occurring [19], in this case predicting the probability of an increase in cigarette consumption during the pandemic, this model predicts outcomes based on selected features, LoR has good performance because it is well-suited for predicting the probability of an event, such as increased cigarette consumption. It uses independent variables to be linearly related to the log odds of the outcome, a relationship between the input variables and the outcome, allowing it to capture the relationship and make accurate predictions. In the context of predicting increased cigarette consumption, LoR’s ability to model the probability of the event contributes to its high precision, recall, and overall performance.

### Strength

This is the first study to investigate the increased number of cigarettes consumed daily by Indonesian smokers during the pandemic using machine learning models. The DT model performed very good, and thus can be used as a tentative reference for predicting the output using the features selected.

### Limitation

There are a few limitations to this work, despite the relative merits of the proposed model compared to the models available in the literature. This paper considered only a few features (27), and more data on economic factors can be incorporated in future research work, as it will enable the real-life application of this model. Another potential limitation of the study is the selection bias introduced by recruiting participants from those who came for vaccination. This sample may not fully represent the general population, as it may primarily include individuals who were willing to get vaccinated lastly, this study was limited by its small dataset, whereas a machine learning model will require large data for training. It is important to acknowledge that these participants may not fully represent the entire smoking population of Indonesia or the broader demographics of kretek smokers. Further experimentation with a large dataset could be done, to test a machine-learning model for the real-time prediction of output.

## Conclusions

The main objective of this study was to develop a modelling framework to predict the increased number of cigarettes consumed daily during the pandemic. The main contribution of this study is in developing a generalized ML-based predictive modelling framework that has been designed to build classification models for predicting the output, based on factors that influenced smoking behaviour during the pandemic. In this study, the DT model achieved the best performance, with an AUC of 98%. Hence, the present work lays the foundation for future research of predicting the increased number of cigarettes consumed based on factors influencing smoking behaviour detailed here. Prediction models can be used to predict smoking cigarette behaviour and inform the authorities about this possibility so that prevention can be suitably carried out. The developed framework can also be used for different healthcare applications and use cases. In this study, an attempt has been made to predict the number of cigarettes consumed during the pandemic based on factors that influence smoking behaviour.

Based on the results of the study, we can conclude that different machine-learning techniques have varying levels of effectiveness in predicting increased cigarette consumption. LoR demonstrated the highest precision and recall values, indicating its prediction accuracy. ANN and DT also performed well in terms of precision and recall. However, NB showed lower precision, accuracy, and sensitivity compared to the other techniques. RF consistently performed well across different metrics. These findings emphasize the importance of considering multiple machine-learning approaches when predicting increased cigarette consumption.

## Data availability statement

Data are available upon reasonable request.

## Conflict of interest statement

We have no conflicts of interest to disclose.

## Funding

None

## Notes

### Competing Interest Statement

The authors have declared no competing interest.

